# Personalized Knowledge-based Graph Neural Networks and Regression Analysis for Computational Diagnosis of High-Risk Cardiovascular Disease Patients

**DOI:** 10.64898/2026.08.24.26361273

**Authors:** Theodosios Lampadarios, Nestoras Karathanasis, Raphael Antartis, Bastian Pfeifer, Dirk von Lewinski, Harald Sourij, George M Spyrou, Anastasis Oulas

## Abstract

Acute myocardial infarction (MI) is a major precursor to heart failure (HF), yet few biomarkers are routinely used to predict post-MI HF, and limited therapeutic options exist to prevent its development. Furthermore, identifying patients at extremely high risk of recurrent MI remains challenging. These gaps highlight the need for improved biomarkers, therapeutic targets, and computational approaches for risk assessment and treatment-response prediction.

To address risk assessment, we developed a systems bioinformatics (SB), graph-based framework representing patient information as personalized networks and integrating omics, clinical, and molecular prior-knowledge data. Graph neural network (GNN) machine learning (ML) models were compared with conventional ML approaches. Two large-scale public plasma proteomic datasets were used to predict post-MI HF. To investigate treatment response, regression models were applied to longitudinal clinical data from >400 hospitalized patients enrolled in the EMMY trial evaluating empagliflozin. ML-driven feature selection identified proteins and clinical parameters with the greatest predictive value.

The graph-based framework demonstrated strong and consistent performance across independent post-MI cohorts. GNN models outperformed conventional approaches, including generalized linear models and XGBoost, particularly when attention mechanisms were incorporated. Using biomarker panels alone, the best GNN achieved an external test AUC of 0.82, compared with 0.77 for the best conventional ML model. When biomarkers were combined with clinical and demographic variables, GNN and conventional ML models achieved AUCs of 0.80 and 0.77, respectively.

Regression models also showed promise for predicting biomarker changes associated with treatment response, with the best model achieving a test RMSE of 0.56. Feature-importance analysis identified NT-proBNP (NPPB), cardiac troponins (TNNI3/TNNT2), and prior HF history as the most influential predictors, consistent with established clinical evidence. Overall, these findings support graph-based ML and regression analysis as promising approaches for improving post-MI HF risk prediction and therapeutic response and identifying clinically relevant markers.

## Introduction

Acute myocardial infarction (MI) is a common predecessor of heart failure (HF) [1,2]. Despite the identification of numerous potential biomarkers, only a limited number have been widely adopted in clinical practice for predicting post-MI HF [2,3]. Adding to this challenge is the lack of novel therapeutic strategies developed in recent years to prevent HF following MI [4]. Furthermore, accurately identifying patients at exceptionally high-risk of experiencing a secondary MI or subsequent HF event remains a significant clinical challenge.

Towards the direction of Predictive, Preventive and Personalized Medicine (PPPM), we propose an intelligent systems bioinformatics (SB) [5] method that harnesses the power of network science to integrate multi-source data (e.g., proteomic, prior knowledge and clinical). This SB approach aims to: 1) Perform high-accuracy, knowledge-based diagnostic prediction of individuals with extremely high-risk of HF post-MI, as well as their response to treatment and 2) Provide an informative set of molecular biomarkers for the task in hand. This work is intended to provide proof-of-concept for the usability of our prediction methodology in clinical practice. These insights can lead to a more effective, early and predictive diagnosis in patients with risk of developing severe illness as well as targeted prevention of therapy for patients at low-risk. A graphical abstract of our risk assessment methodology as well as its potential integration in a clinical/diagnostic setting is shown in Figure 1.

**Figure 1.**
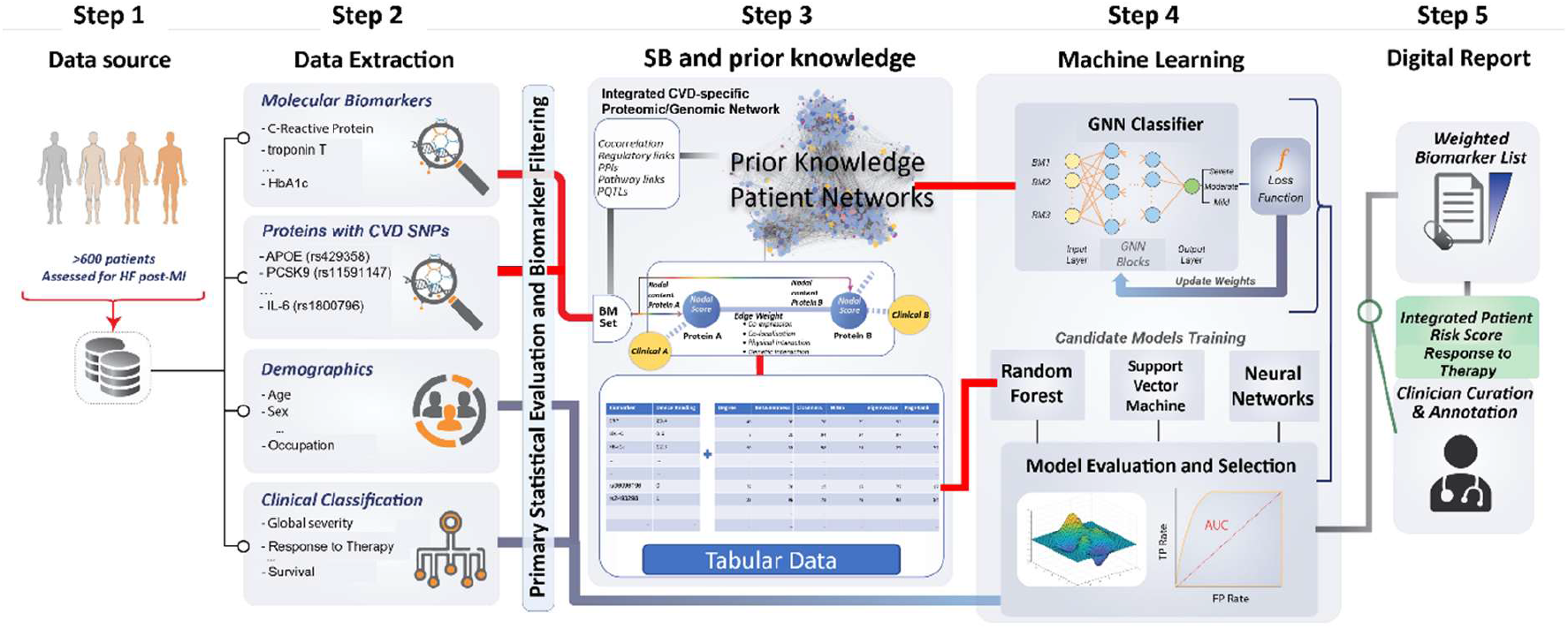
Graphical abstract of our SB approach and potential integration in a clinical setting. **Steps 1 -** Data sources, **Step 2** – Data extraction, **Step 3** – Systems Bioinformatics (SB) biomarker integration with prior knowledge, **Step 4 -** Machine learning (ML) algorithms, Graph Neural Network (GNNs) and other ML tools. **Step 5 –** Potential integration with clinical setting via Digital Reporting and clinician curation.

A key feature of our methodology is the integration of multi-level biological and clinical information derived from diverse data sources. Leveraging prior biological knowledge and systems biology tools such as GeneMANIA, we first constructed a backbone cardiovascular network by integrating multiple layers of evidence. This network incorporated established cardiovascular disease (CVD) biomarkers, including NT-proBNP (NPPB), cardiac troponin I (TNNI3), cardiac troponin T (TNNT2), C-reactive protein (CRP), and glycated hemoglobin (HbA1c), together with genes containing genetic variants previously associated with cardiovascular disease. Networks obtained from the STRING database were subsequently integrated using proteins measured in the dataset and subsets of the selected biomarkers. Using two large-scale, publicly available post-myocardial infarction (post-MI) proteomic cohorts, we then overlaid patient-specific proteomic and clinical data onto this knowledge-driven network framework. Additional prior knowledge, including gene–disease associations (GDAs) and Gene Ontology (GO) annotations, was incorporated to further enrich the personalized patient networks. Within these networks, nodes encoded biomarker expression levels, protein identity, GDA information, GDA-weighted expression values, and GO term–gene mappings, while edges represented relationships and similarities among these features. This approach transformed heterogeneous, multi-source data into a unified graph-based representation suitable for downstream machine learning analyses.

Given the graph-structured nature of the resulting patient representations, we employed graph neural networks (GNNs) to develop predictive models for heart failure (HF) following MI. To benchmark performance, several conventional machine learning approaches were also evaluated. Our results demonstrate that GNN-based models, particularly those incorporating attention mechanisms, consistently outperformed traditional machine learning methods, including generalized linear models (GLMs) and XGBoost, as well as the clinically established GRACE score for HF prediction. Furthermore, GNN models utilizing selected biomarker panels, either alone or in combination with clinical and demographic variables, achieved the strongest generalization performance on an independent external test cohort across all evaluated feature combinations.

The incorporation of attention mechanisms enabled the models to assign differential importance to protein interactions and network components, thereby highlighting features with greater diagnostic and prognostic relevance. Notably, NT-proBNP (NPPB), cardiac troponins (TNNI3/TNNT2), and a prior history of heart failure consistently emerged among the most influential predictors across ensemble machine learning models, reinforcing their well-established clinical significance. In addition, Kaplan–Meier time-to-event analyses independently validated the prognostic importance of several identified features associated with HF development, further supporting the biological and clinical relevance of the predictive signals captured by the proposed framework.

The EMMY trial dataset [6], in which patients were randomised to either the drug Empagliflozin or placebo for 26 weeks, offers a unique longitudinal framework not captured in the two previously described proteomic datasets. The availability of repeated clinical visits, patient histories, and serial laboratory measurements enables a more comprehensive assessment of treatment response and supports the potential for earlier therapeutic intervention. In this cardiovascular high-risk cohort, Empagliflozin treatment was associated with an improvement in the primary outcome measure, change in NT-proBNP level, compared with placebo. Our regression model, which integrates longitudinal data, biomarker profiles, and clinical features, predicts NT-proBNP levels at visit 4 (26 weeks after treatment initiation), and may therefore facilitate earlier identification of therapeutic response and provide an additional risk stratification tool prior to clinically critical time points.

## Methods

### Overall Risk Assessment Methodology

Patient data were derived from three public datasets for CVD [2]. Two of these included proteomics data for patients assessed for HF post-MI. This data was used to obtain: 1) Protein expression values for four widely accepted CVD biomarkers (haemoglobin A1 (HbA1), C-reactive protein (CRP), N-terminal pro-brain natriuretic peptide (NT-proBNP/NPPB) and high-sensitive troponin T and I (hsTnT/TNNT2 and hsTnI/TNNI3)) [2–4,6]. 2) Protein expression values for eight genes which are known to contain variants previously implicated in CVD and 3) Patient clinical and demographic information. The third dataset included molecular laboratory experiments for the four biomarkers listed above in addition to clinical and patient history data. This clinical trial dataset (EMMY) is comprised longitudinal data (from 4 patient visits) and was used to observe response to treatment over time for the drug empagliflozin. Results showed that observing the NT-proBNP change over time was the primary outcome in predicting response to treatment (see Error! Reference source not found.**1 – Steps 1 and 2** - Data source and extraction). In addition to personalized patient data, we further included prior knowledge in the form of information from public databases to generate integrated knowledge-based systemic networks. This was achieved using the GeneMANIA tool (https://genemania.org/) [7]. The tool was used to create connections between the CVD-related proteins/biomarkers described above and generate networks using various public repositories (e.g., BioGRID [8]). These networks incorporate information for protein-protein interactions, gene-to-gene relationships, common pathways involved and co-expression values. This SB approach consequently boosts the feature set available for CVD biomarkers with network topological measures (degree, betweenness, closeness, etc.) thus transforming and potentially enhancing the input that will be passed downstream to ML model(s) (**see** Error! Reference source not found.**1 - Step 3** - Biomarker integration with prior knowledge). Next, we performed benchmarking for a wide range of ML algorithms using standard training, validation and testing procedures. To accommodate for the SB approach adopted herein, our primary choice was the use of Graph Neural Networks (GNNs). Each patient was represented as a personalized graph incorporating multiple levels of information. The protein biomarkers were depicted as nodes containing expression values as embeddings. Genomic information was added indirectly as nodes in the network (due to missing genotypic information). Specifically, proteins were selected based on evidence supporting their role as protein quantitative trait loci (pQTLs) associated with cardiovascular disease (CVD)–related variants (see Table 1). These protein expressions are directly affected by the CVD variants; therefore, their expression values were also added on the corresponding nodes on the networks. In addition, interactions between biomarkers were depicted as edges containing an encoding depending on the difference in expression between nodes. These personalized patient graphs were used to train GNNs in order to predict risk for HF post-MI for the two proteomic datasets. Additional more traditional ML classifiers in the form of generalized linear models (GLMs), decision trees (XGBoost) and other deep Artificial Neural Networks (ANNs) were employed using tabular data to obtain prediction accuracy comparisons and ultimately choose the best classifier for the job. Due to the lack of proteomic data to generate networks, the EMMY dataset was analyzed using the latter ML approaches applied on tabular data in order to predict response to treatment (see Error! Reference source not found.**1 - Step 4 machine learning**). Finally, we aspire that our approach can potentially be applied in a clinical setting in the future. This will entail **model explainability and reporting** utilizing the output of the GNN and other ML algorithm(s) to deliver a risk score, as well as importance scores for a protein biomarkers and clinical variables (weighted biomarker list). Thus, providing and optimal patient stratification and response to treatment prediction (see Error! Reference source not found.**1 - Step 5 Digital Report**).

**Table 1.**
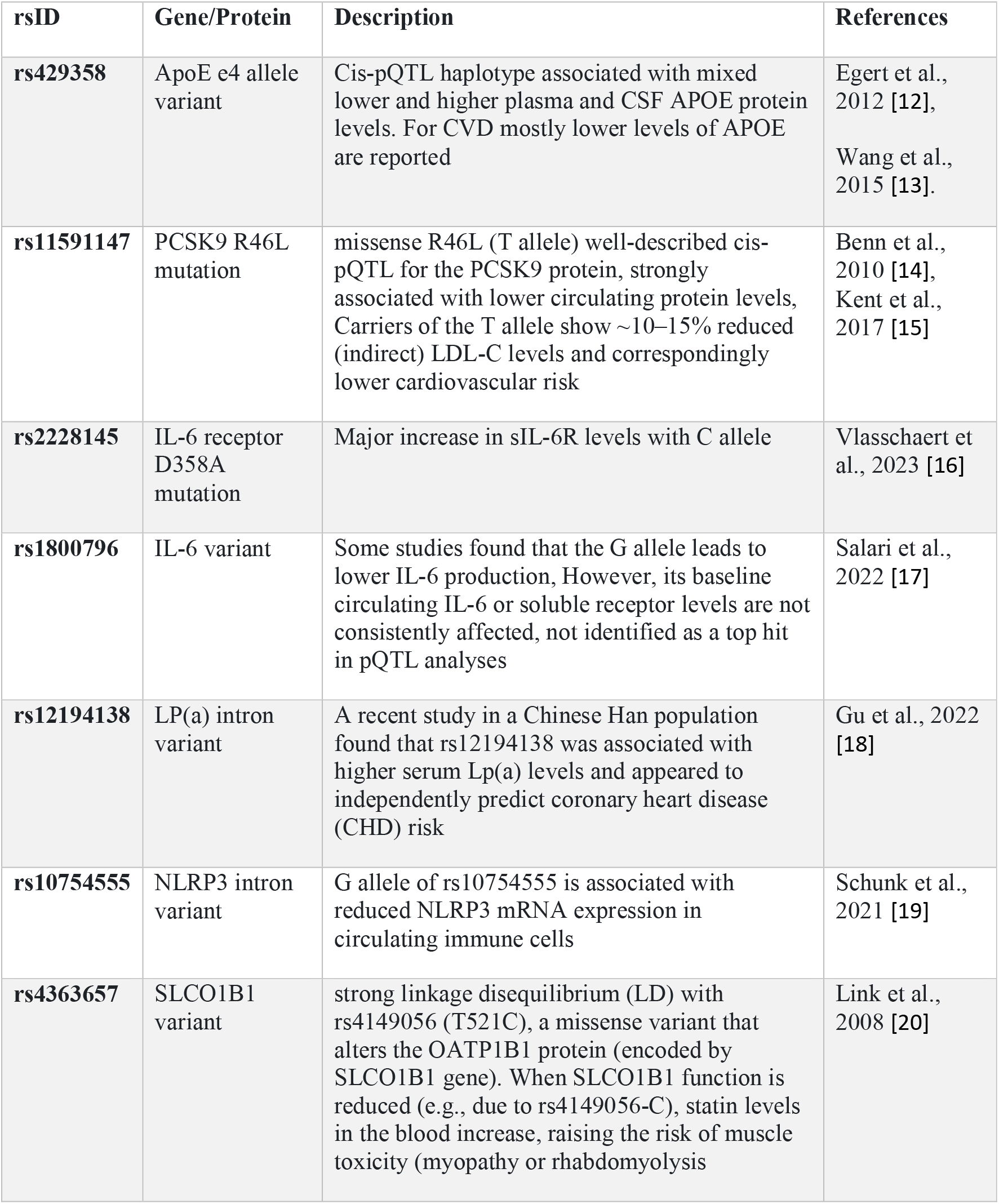
Genes containing variants implicated in CVD.

### Datasets

A publicly available dataset was utilized that specifically addresses the question described above. Namely, stratification of CVD patients with prior myocardial infarction (MI) that are at extremely high-risk of subsequently experiencing a secondary event (i.e., heart failure (HF)).

The dataset is comprised of two cohorts. Both used aptamer-based affinity-capture plasma proteomics to measure plasma proteins at post-MI [2]:

New Zealand cohort (CDCS - Coronary Disease Cohort Study) - 431 post-MI patients, of which **181** who were subsequently hospitalized for HF in comparison with **250** patients who remained event free over a **median follow-up of 4.9 years**

Singapore cohort (IMMACULATE [Improving Outcomes in Myocardial Infarction through Reversal of Cardiac Remodelling]) - 223 patients post-MI, of which **33** patients were hospitalized for HF (**median follow-up, 2.0 years**).

The EMMY trial cohort is comprised of 476 hospitalized patients [6] with acute myocardial infarction accompanied by a large creatine kinase elevation (>800 IU/L). These patients were assigned to empagliflozin 10 mg or matching placebo once daily within 72 h of percutaneous coronary intervention. The EMMY trial data is comprised of clinical and demographic data (for details see supplementary information). It was used in order to assess the response to treatment for the drug empagliflozin on already high-risk CVD patients for follow-up visits scheduled at 6, 12, and 26 weeks after first drug administration. This was achieved by observing the N-terminal pro-hormone of brain natriuretic peptide (NTproBNP) change over time as the primary outcome.

### Backbone Biomarker Network

A backbone network was created using four established biomarkers commonly used in CVD for cardiovascular risk assessment and stratification, selected in consultation with clinicians. These include HbA1c, CRP, NT-proBNP/NPPB and TnT/TNNT2. In addition, six genes which are known to contain variants previously implicated in CVD where also included (see Table 1). The latter six SNP-containing genes were selected by scanning public databases, including dbSNP [9] and pQTL databases [10], which catalog genetic variants and link polymorphisms to protein biomarkers. Specifically, proteins were selected based on evidence supporting their role as protein quantitative trait loci (pQTLs) associated with cardiovascular disease (CVD)–related variants. Therefore, their expression is directly affected by their SNPs. All ten genes were used as input to the SB tool GeneMANIA and a densely connected network was obtained using prior-knowledge information retrieved from online database utilized by the tool, including additional neighboring proteins (see Figure 2). These include databases like BIOGRID, and other publicly available data which provide molecular information with respect to protein-protein interactions, gene-to-gene relationships, common pathways involved, co-expression and co-localization.

**Figure 2.**
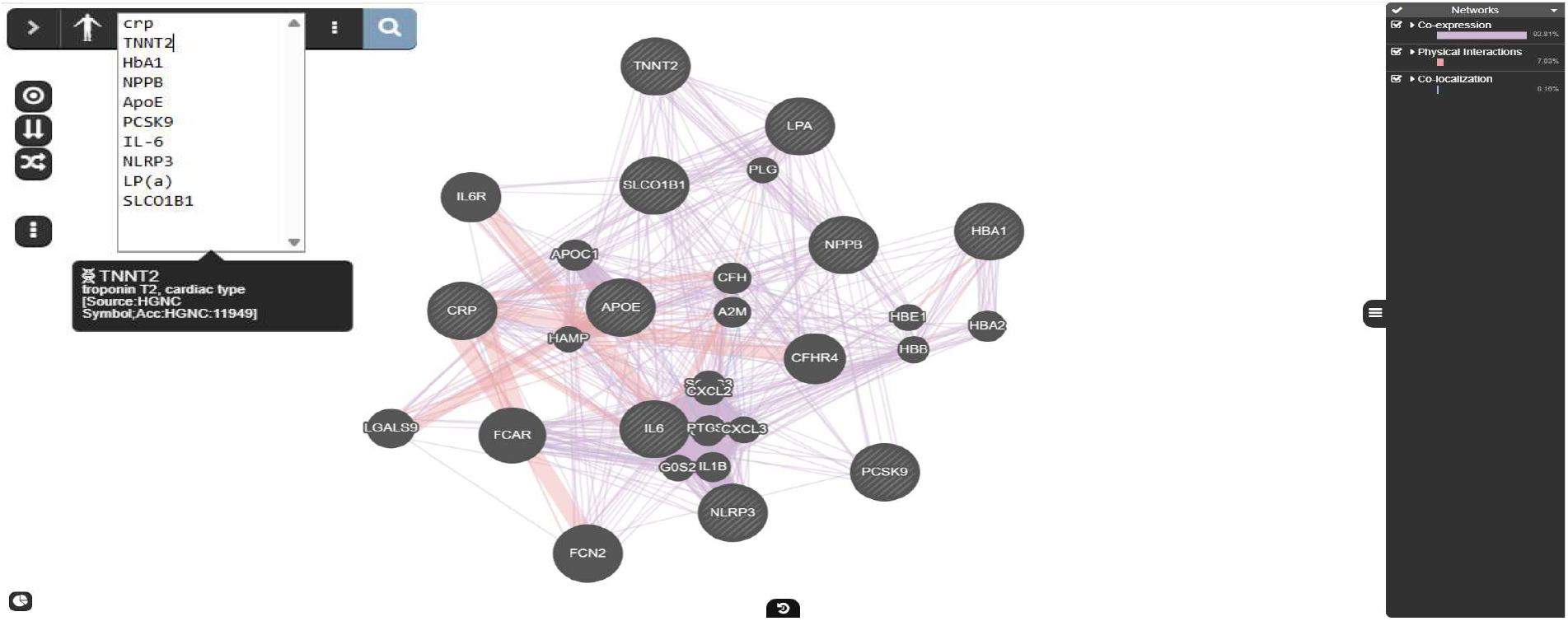
GeneMANIA snap-shot of backbone prior-knowledge CVD network. It contains biomarkers and genes commonly implicated in CVD as well as neighboring genes linked by molecular information.

By default, GeneMANIA has tendance to introduce neighboring proteins, not present in the original input list. Subsequently, to establish a framework for training and testing our GNN models, we first identified the common proteins between the GeneMANIA network and the proteomics dataset, and then used the STRING database [11] to construct interaction networks which exclusively contain these proteins. Interaction networks were generated using the complete set of common proteins or selected subsets, using a minimum interaction score of 0.4 for the edges. The threshold was adjusted to avoid a disconnected networks. Specifically, we lowered the score and selected the highest value that yielded a connected network. In cases of multiple edges between nodes, only a single edge was retained.

### Data Preprocessing

#### CDCS – IMMACULATE datasets –

Several preprocessing steps were essential to prepare the data for ML algorithm(s). From the CDCS dataset, patients were included if they were categorized as Control, HF_First or HF_and_MI_together according to the Somalogic Group Full variable (for details see supplementary information). In the Immaculate dataset, inclusion was restricted to patients with non-missing values for the HF variable. Additionally, proteins that had not passed the quality control and variables with more than 20% of missing values or with no variability (i.e., single unique value) were removed. Subsequently, mean and mode imputation was applied to handle missing values in continuous and categorical variables, respectively. A log2-transformation was applied to protein expression data as well as NT-proBNP and hsTnI values from the clinical data, followed by z-score normalization on both protein expression and numerical clinical data. Categorical variables were one-hot encoded either automatically within the training framework when required by specific models, or using the OneHotEncoder from scikit-learn [21] library.

#### EMMY dataset

***-*** From the EMMY dataset, only patients who received the drug (empagliflozin) and had non-missing NT-proBNP values at visit 4 were included. In addition, NT-proBNP measured at the central lab for visits 1,2 and 4, along with the local lab measurement at visit 3 (the only available measurement for that visit), were retained. BMI and change of left-ventricular ejection fraction (LVEF) were also calculated for visits 2 and 4. Variables with more than 20% of missing values or with no variability (i.e., single unique value) were removed. Subsequently, mean imputation for continuous variables and mode imputation for categorical variables to handle missing data. A log_2_-transformation was applied to NT-proBNP, TnT, hsTnT, HbA1 as well as CRP, followed by z-score normalization of all numeric variables. Categorical variables were automatically one-hot encoded within the training framework for models that required it.

### Creating Personalized Patient Networks Enriched with Prior-Knowledge

The backbone networks described above are generic networks derived from prior knowledge (i.e., molecular information). Subsequently, we created personalized patient networks by incorporating specific patient information and protein expression values. These personalized patient networks were further enriched with Gene-Disease Association (GDA) scores for heart failure obtained from DISGENET [22], for each corresponding protein. When no GDA was available for a given protein, 0 was assigned. An additional node feature obtained by multiplying the expression value with the GDA score was also included. Moreover, nodes features were extended with two distinct one-hot-encoded representations, a Gene Ontology (GO) term – gene mapping and another encoding for protein identity. Specifically, the proteins from GeneMANIA network were used as input to g:Profiler (accessed on 16 April 2026, Ensembl 113) [23] to identify associated GO terms. This produced a matrix indicating the presence or absence of an association between each protein and each GO term. The second encoding was applied to represent individual proteins uniquely within each network (see Figure 3).

**Figure 3.**
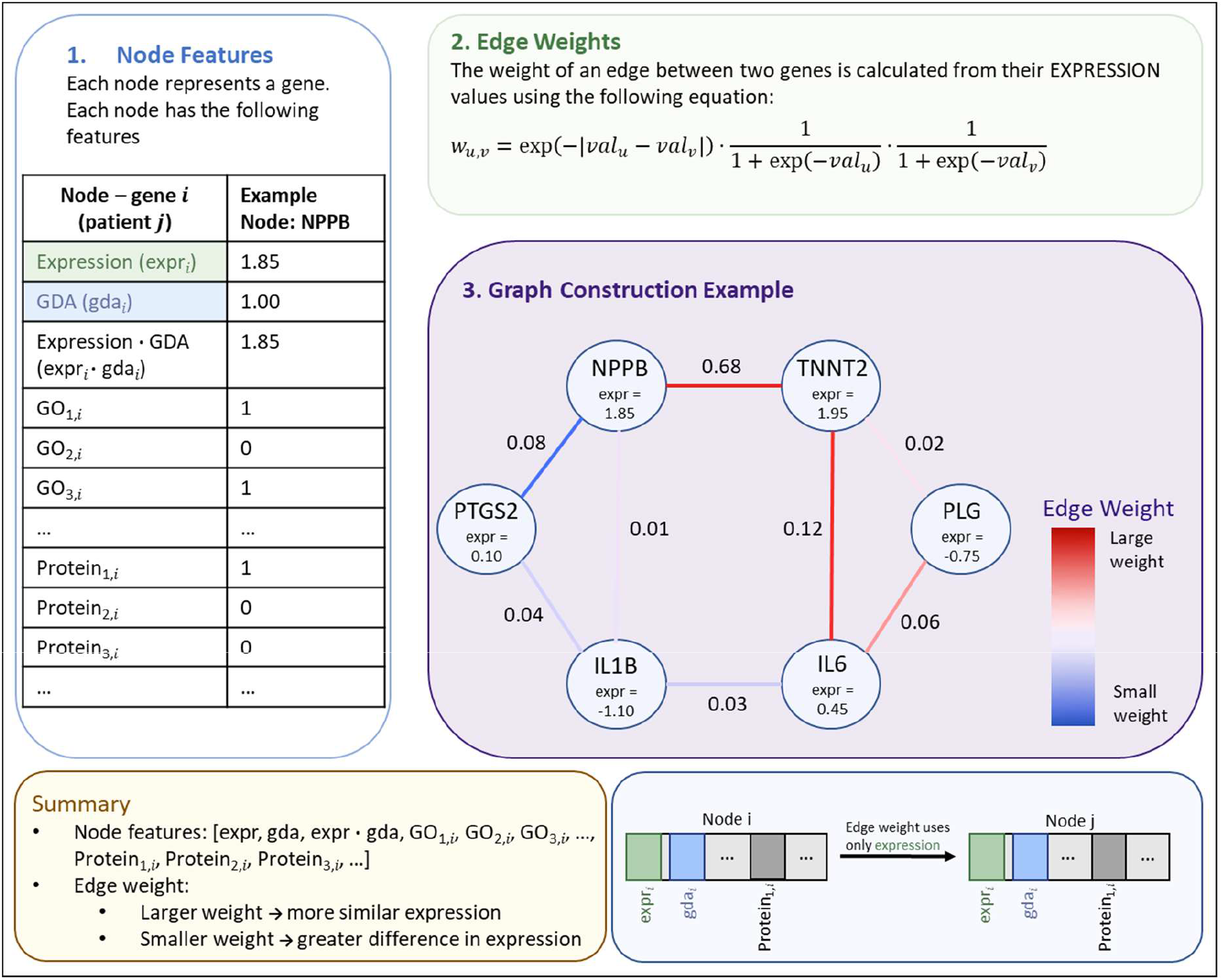
Graph construction and node feature representation. Each node in the graph represents a gene for a given patient and is associated with a feature vector that contains an expression value, a GDA score, expression value ⋅ GDA, Gene Ontology (GO) term – gene mapping and one-hot encoding for protein identity. For example, NPPB gene is mapped to GO_1,_*_i_*, GO_3,_*_i_* and Protein_1,_*_i_*. Edge weights between connected genes are computed from their expression values using the defined similarity function, where genes with more similar expression profiles receive larger edge weights. The graph construction example presents how node expression values determine edge strengths, visualized by the edge color gradient.

In addition, the edge weights were also modified to reflect personalized patient information. This was achieved using the following equation that captures both the similarity in expression levels between to nodes as well as the individual expression of each node:

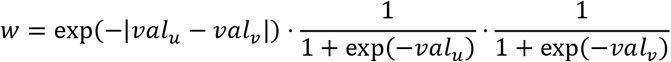

Where *val_u_* is the expression value of node *u* and *val_v_* is the expression value of node *v*. The weighting formula assigns high weights to node pairs that are both similarly and highly expressed and low weights to pairs that are either lowly expressed or exhibit opposing expression patterns.

### Training, Validation and Testing of GNN and other ML models

#### Deep-learning Graph Neural Network Models

For the best-performing combinations with backbone network-selected proteins in ML models, we developed GNN models to compare performance. Networks were split into training and test sets and the training and test performance was recorded. The training/validation procedure was performed using a stratified 5-fold nested cross-validation procedure. GNNs were trained on personalized patient networks using the PyTorch library [24] on GPU NVIDIA A30 setup. We developed two separated architectures one based on multiple stacked GCNConv layers and the other on multiple stacked layers GATv2Conv, followed by GraphNorm, LeakyReLU activation and dropout regularization. GATv2Conv also includes an internal dropout mechanism, in addition to the standard dropout layers used for regularization. Node embeddings were aggregated into graph-level representations using both global mean and sum pooling, which were concatenated to form the final graph embedding. In cases where protein expression data were combined with demographic (e.g., age) and clinical information (e.g., History of Hypertension (HHT)), these features were concatenated with the graph embeddings and used as input to a Multi-layer Perceptron Classifier (MLP) for final prediction. When such additional data were not available, only the graph embeddings were used as input to the MLP.

Additionally, we applied grid search for the optimization of the GNNs. Area Under the Curve (AUC) metric were used to monitor training and validation. Early stopping was also applied based on validation AUC with a patience of 10 epochs. The total number of training epochs was determined as the mean number of epochs across CV folds. Furthermore, whether the calculated weight was used or not was determined through grid search. Model performance was evaluated using 5-fold nested cross-validation (CV) and the results were averaged across folds (see Figure 4). The optimal set of hyperparameters was selected based on the nested CV and subsequently applied to the test set.

**Figure 4.**
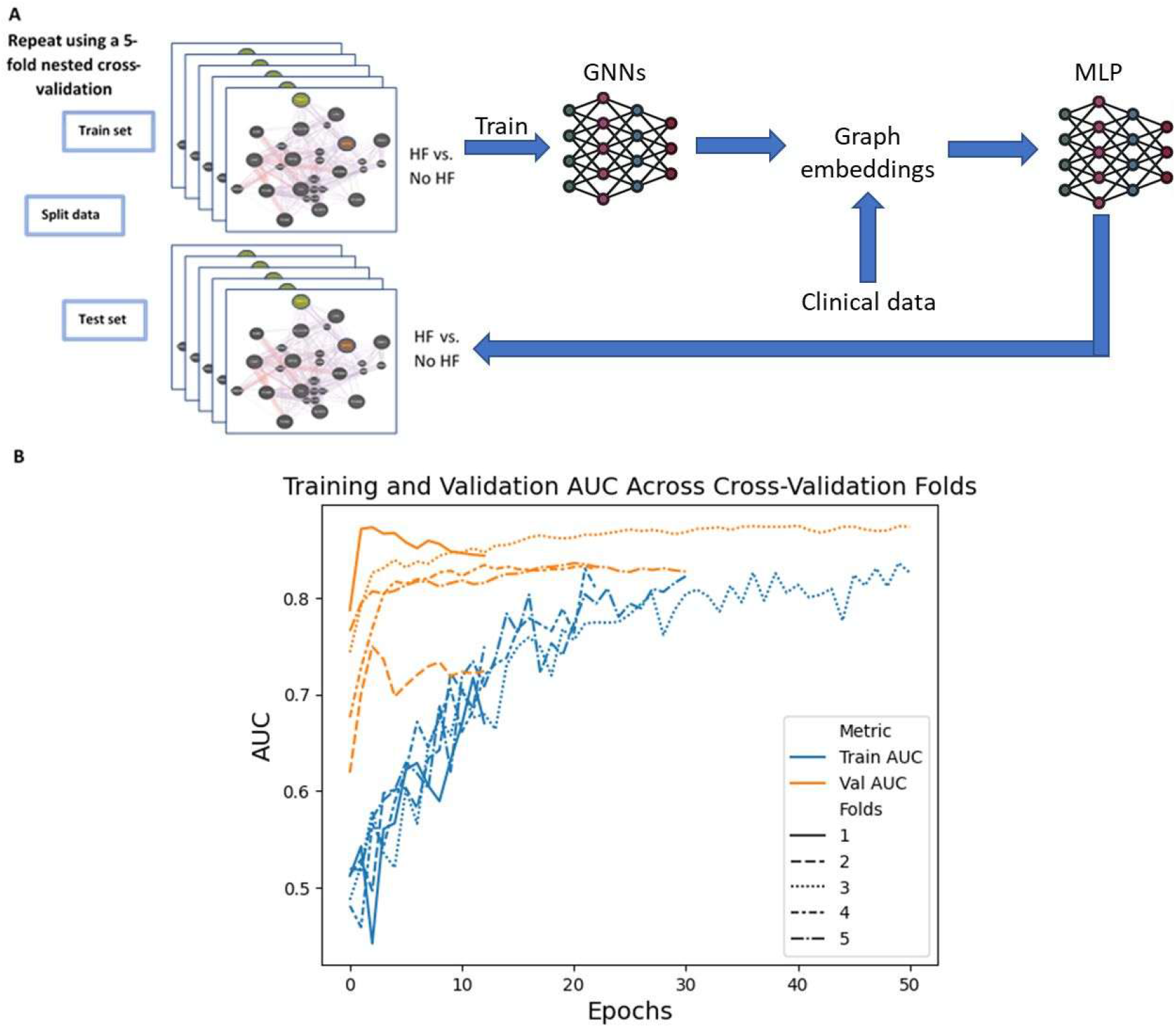
Training/validation and procedure of GNN models. **A.** Flowchart describing the randomized split of the data into training, validation and test sets in order to predict patients who suffered HF post-MI. **B.** Typical GNN learning curves across CV folds.

#### Other ML algorithms

Additional ML algorithms were employed to predict HF post-MI. The H2O [24] R package was utilized, providing an ensemble of tools that can be used for comparison with our GNN approach for patient classification. Specifically, H2O AutoML was used to build up to 30 models, from which stacked ensemble models were subsequently developed. Similar to the GNN approach, a stratified 5-fold nested cross-validation procedure was adopted and AUC was used to evaluate the models. All tools used can be summarized in Table 2.

**Table 2.** Description of GNN and other ML tools.

| Acronym | Full Name |
| --- | --- |
| GCNConv | Graph Neural Network |
| GATv2Conv | Graph Neural Network with Attention learning |
| DeepLearning | Deep Neural Network |
| DRF | Distributed Random Forest |
| XRT | Extremely Randomized Trees |
| GBM | Gradient Boosting Machine |
| GLM | Generalized Linear Model |
| **XGBoost** | Extreme Gradient Boosting |
| **StackedEnsemble** | Stacked Ensembles |

For the EMMY data, the task in hand requires a **regression ML** analysis, as the aim is to predict the NT-proBNP value, which according to the EMMY trial study, is the primary outcome that changes between treatment and placebo groups over time [6]. As described above, a similar stratified 5-fold nested cross-validation procedure was applied. Under these circumstances different metrics are required to assess our model performance, namely we used Root Mean Squared Error (RMSE) as the primary evaluation metric and Mean Absolute Error (MAE) as an additional measure (see Figure 5).

**Figure 5.**
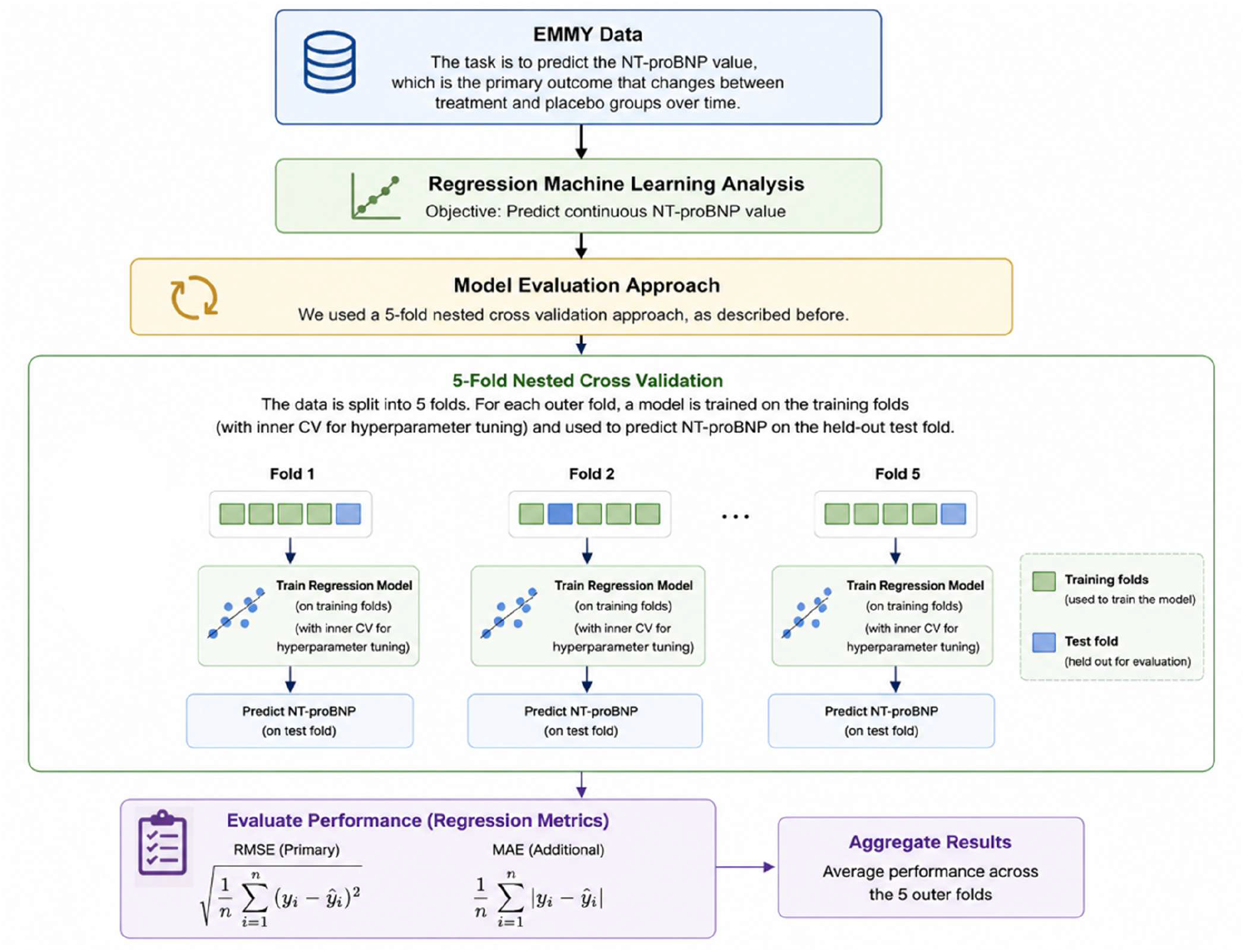
Flowchart of the EMMY data analysis. The chart shows the regression model training and cross validation process and the prediction of NT-proBNP at patient visit 4, 26 weeks after drug (empagliflozin) administration.

### Feature Selection Methods

CDCS and IMMACULATE datasets include some SOMAmers reagents (definition in supplementary information) that target the same proteins. Therefore, Ridge Regression via H2O was applied to identify the most informative reagent for each protein, ensuring that a single protein was used when mapping each node in the corresponding network.

In addition, feature selection methods were employed to improve the performance of ML models. Specifically, a LASSO regression [25,26] model was implemented using the H2O package to identify the most significant features. LASSO is a regularization method that applies an L1 penalty to the regression coefficients, which promotes sparsity in the coefficient vector by shrinking less important coefficients to zero and thus selecting a set of important features.

Moreover, a type of forward selection approach was also applied to identify the features that yield the best model performance. Specifically, feature importance scores from each outer training set were used to rank the features, which then iteratively were added, one at a time, following that ranking and the feature set that achieved the best performance was retained, up to a maximum of 100 features or the total number of available features, whichever was lower.

## Results

### ML Feature Importance

For ML tools implemented via H2O, a feature importance analysis was performed, which allows for a ranking of all features as considered across the best explainable models used in the analysis of each case. Some models by default allow for feature elimination by coefficient values generated during the training process per se. The LASSO algorithm has an in-built ranking procedure that allows for such a feature prioritization and can thus be used to rank features according to the model-generated coefficients. In addition, we performed a type of forward selection approach whereby we incrementally increased the number of top ranked features until a model performance plateau is achieved. For the CDCS dataset, the best performing combinations based on mean CV AUC were achieved using features selected by LASSO from combined expression and clinical features, as well as using forward selection and LASSO on both combined and expression-only feature sets (see Table 5). The highest-ranking features in the combined data with LASSO and forward selection included LTA4H, hsTnI, H2AFZ, NT-proBNP and others (see Figure 6A, B, C). As for the EMMY dataset, the best performing combinations based on mean CV RMSE were those including features from visits 1, 2 and 3 selected by only LASSO as well as selected by both LASSO and forward selection. In both cases, the highest-ranking features included NTproBNP from visits 2 and 3, LVEF from visit 1 and others (see Figure 6D, Supplementary Figure S1).

**Figure 6.**
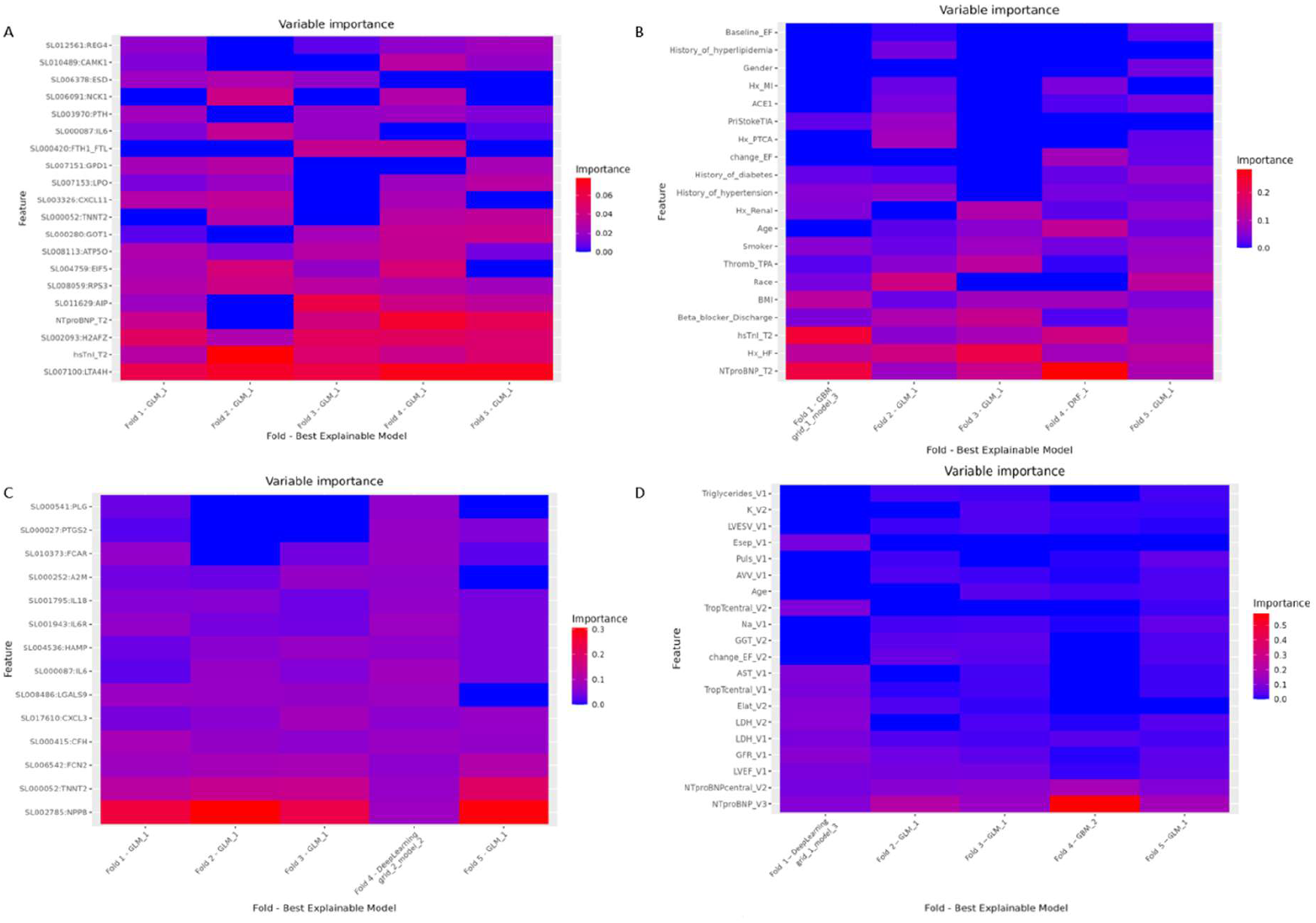
Feature importance heatmaps. **A.** CDCS data top 20 features derived from proteomic and clinical data, selected by LASSO and forward selection, as evaluated by the best explainable H2O model across each fold. **B.** CDCS data top 20 features derived from clinical data, selected by LASSO and forward selection, as evaluated by the best explainable H2O model across each fold. **C.** CDCS proteomics data with selected proteins from backbone network and LASSO, as evaluated by the best explainable H2O model across each fold. **D.** EMMY trial data top 20 features derived from Visits 1,2 and 3, selected by LASSO, as evaluated by the best explainable H2O model across each fold.

### Functional enrichment analysis

In the node features, we included a GO term – gene mapping generated using the GeneMANIA network proteins in g:Profiler [23]. This analysis was performed for both the CDCS and IMMACULATE datasets (see Table 3 and Supplementary Table S1 respectively). The enrichment analysis revealed significant involvement of inflammatory and immune-related processes and tissue remodeling processes. Immune related terms, such as signaling receptor binding, neutrophil chemotaxis and interleukin-6 receptor complex indicate activation of inflammatory pathways known to drive cardiac dysfunction [27,28]. In addition, extracellular matrix and fibrosis-related processes, including collagen-containing extracellular matrix and negative regulation of collagen biosynthetic process suggest ongoing structural remodeling of cardiac tissue [29]. Vascular related pathways, such as circulatory system development, further support the involvement of altered vascular function [30].

**Table 3.** Results from functional enrichment analysis of 15 GeneMANIA network proteins for CDCS.

| Term ID | Term Name | Adjusted p-value |
| --- | --- | --- |
| GO:0005102 | signaling receptor binding | $3.067 \times 10^{-6}$ |
| GO:0045236 | CXCR chemokine receptor binding | $2.027 \times 10^{-2}$ |
| **GO:0044419** | biological process involved in interspecies interaction between organisms | $1.141 \times 10^{-9}$ |
| **GO:0051241** | negative regulation of multicellular organismal process | $3.124 \times 10^{-4}$ |
| **GO:0002384** | hepatic immune response | $6.419 \times 10^{-4}$ |
| **GO:0072359** | circulatory system development | $8.240 \times 10^{-3}$ |
| **GO:0042592** | homeostatic process | $1.089 \times 10^{-2}$ |
| **GO:0031394** | positive regulation of prostaglandin biosynthetic process | $1.345 \times 10^{-2}$ |
| **GO:0001781** | neutrophil apoptotic process | $1.793 \times 10^{-2}$ |
| **GO:0032966** | negative regulation of collagen biosynthetic process | $2.304 \times 10^{-2}$ |
| **GO:0002294** | CD4-positive, alpha-beta T cell differentiation involved in immune response | $3.524 \times 10^{-2}$ |
| **GO:0030593** | neutrophil chemotaxis | $3.785 \times 10^{-2}$ |
| **GO:0048661** | positive regulation of smooth muscle cell proliferation | $3.920 \times 10^{-2}$ |
| **GO:0005576** | extracellular region | $4.717 \times 10^{-6}$ |
| **GO:0005896** | interleukin-6 receptor complex | $1.768 \times 10^{-4}$ |
| **GO:1905370** | serine-type endopeptidase complex | $8.965 \times 10^{-3}$ |
| **GO:0062023** | collagen-containing extracellular matrix | $1.634 \times 10^{-2}$ |

### GNN Optimization

GNNs were optimized using a grid-search approach, whereby all avaibale hyperparameter combinations were assessed based on AUC valued derived from the training/validation procedure for our models. The full list of hyperparameters for our GATv2Conv models is shown in Table 4. Similar hyperparameters were also assessed for the GCNConv models (see Supplementary Table S2)

**Table 4.** Hyperparameter search space used for grid search optimization of the GATv2Conv model.

| Parameters | Hyperparameter search space |
| --- | --- |
| **Hidden channels** | [32, 64] |
| **Number of GATv2Conv layers** | [1, 2] |
| **Heads** | [2, 4] |
| **Number of MLP layers** | [1, 2] |
| **GAT dropout** | [0.1, 0.3, 0.5] |
| **MLP dropout** | [0.1, 0.3, 0.5] |
| **Learning rate** | [0.001] |
| **Epochs** | [200] |
| **Use of custom edge weights** | [True, False] |

### Comparison of GNN and other ML tools for HF prediction post-MI

In order to perform a comprehensive comparison of multiple ML tools for the different tasks in hand, we assessed an ensemble of ML tools as well as different graph neural network algorithms (see Table 2). In addition, we evaluated a wide range of features in our classification schemes with the intent to obtain the optimal set of features for the ML task. Finally, we report the performance achieved by the best model in each of the validation/test procedures (for the CDCS dataset see Table 5). We observed that the combination of expression and clinical/demographic data with forward selection and without feature selection, achieved optimal test performance (AUC: 0.92). In addition, the combination of expression and clinical/demographic data achieved the best CV performance (AUC: 0.98) after the application of LASSO alone or in combination with forward selection. Also, expression data in combination with forward selection and LASSO yielded AUC: 0.98. Furthermore, GNN models yielded higher AUC performance in comparison with ML models on the same combination of data in both CV and Test (see Table 5, Figure 7, supplementary Figures S2 and S3).

**Figure 7.**
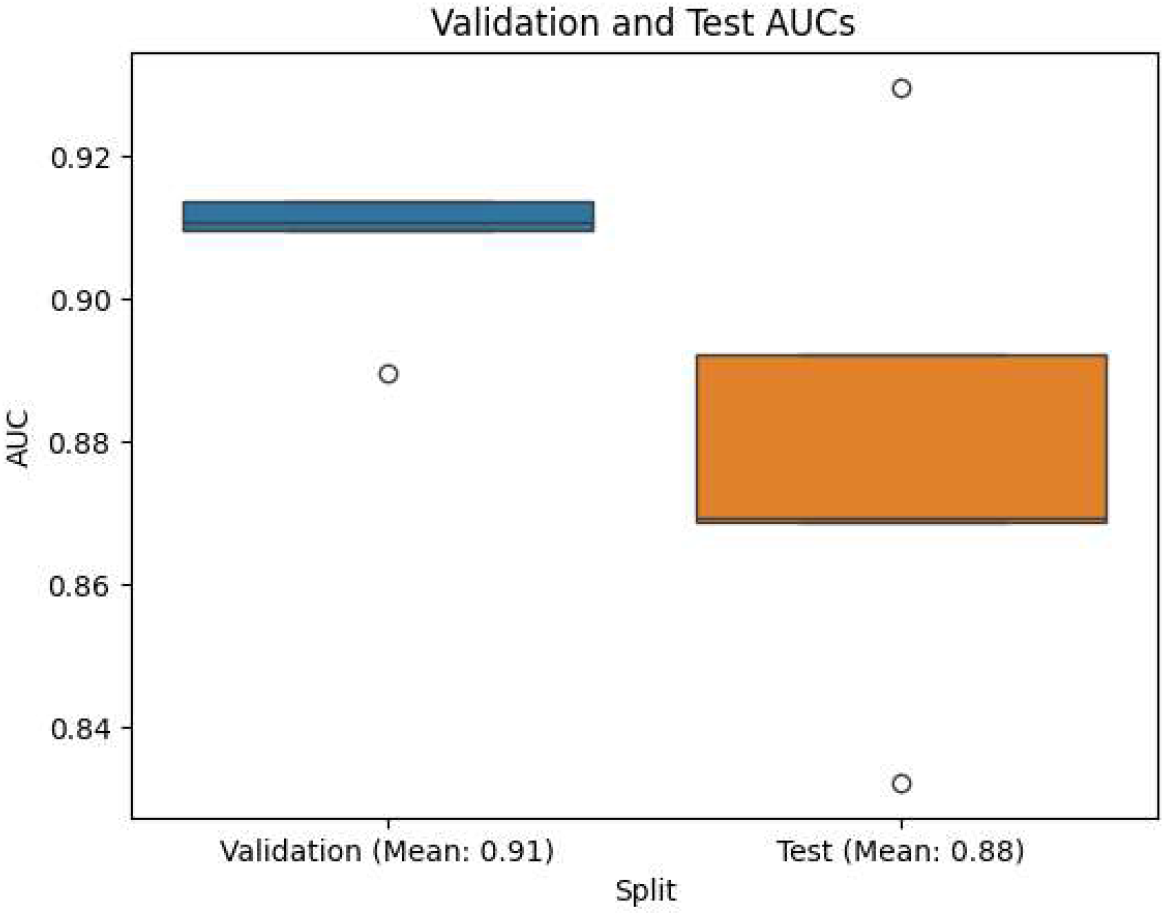
GATv2Conv performance box plot displaying 5-fold nested cross-validation runs on network combined data with features selected by FS and LASSO.

**Table 5.** Combination of features vs. classification performance AUC on CDCS dataset, sorted by mean Test AUC (exprs: expression data; clinical: clinical data; combined: expression and clinical data; net_exprs: expression only for proteins from the backbone network; net_combined: expression only for proteins from the backbone network combined with clinical/demographic data; fs: forward selection approach; lasso: LASSO selection). Note: net_combined with FS-selected features was excluded from GNN models because the selected nodes formed a disconnected network.

| Combinations | lasso | fs | Validation (Mean AUC) | Test (Mean AUC) |
| --- | --- | --- | --- | --- |
| combined (other ML) |  | ✓ | 0.95 | 0.92 |
| combined (other ML) |  |  | 0.91 | 0.92 |
| combined (other ML) | ✓ |  | 0.98 | 0.91 |
| exprs (other ML) |  |  | 0.89 | 0.91 |
| exprs (other ML) |  | ✓ | 0.94 | 0.90 |
| combined (other ML) | ✓ | ✓ | 0.98 | 0.89 |
| exprs (other ML) | ✓ | ✓ | 0.98 | 0.89 |
| exprs (other ML) | ✓ |  | 0.97 | 0.89 |
| net_combined (GATv2Conv) |  |  | 0.90 | 0.89 |
| net_combined (GCNConv) | ✓ | ✓ | 0.91 | 0.88 |
| net_combined (GATv2Conv) | ✓ | ✓ | 0.91 | 0.88 |
| net_combined (GATv2Conv) | ✓ |  | 0.91 | 0.88 |
| net_combined (GCNConv) |  |  | 0.90 | 0.88 |
| net_combined (GCNConv) | ✓ |  | 0.90 | 0.88 |
| net_combined (other ML) |  | ✓ | 0.89 | 0.88 |
| net_combined (other ML) | ✓ |  | 0.89 | 0.88 |
| net_combined (other ML) |  |  | 0.89 | 0.88 |
| net_combined (other ML) | ✓ | ✓ | 0.89 | 0.87 |
| clinical (other ML) | ✓ | ✓ | 0.89 | 0.87 |
| clinical (other ML) |  |  | 0.88 | 0.87 |
| clinical (other ML) | ✓ |  | 0.89 | 0.86 |
| clinical (other ML) |  | ✓ | 0.89 | 0.86 |
| net_exprs (other ML) | ✓ |  | 0.82 | 0.81 |
| net_exprs (other ML) | ✓ | ✓ | 0.82 | 0.79 |
| net_exprs (other ML) |  | ✓ | 0.82 | 0.78 |
| net_exprs (other ML) |  |  | 0.82 | 0.78 |

Next, the same approaches were applied to the full CDCS dataset to train the models, which were evaluated on the IMMACULATE dataset as an external test cohort. Feature combinations with the highest CV AUC performance failed to generalize, showing significantly lower AUC performance on the test dataset. In contrast, feature combinations based on network protein expression alone or combined with clinical/demographic data, including the full set as well as subsets selected by using forward selection or both forward selection and LASSO, achieved lower validation AUC but higher test AUC than the other combinations, indicating better generalization. For example, on the combination of network protein expression data we observed that GATv2Conv outperformed ML models. Specifically, GATv2Conv yielded AUC 0.83 in CV and 0.82 in test, in comparison with ML models that had 0.82 in CV and 0.77 in test. (see Table 6).

**Table 6.** Combination of features vs. classification performance AUC, with training on CDCS dataset and external test on IMMACULATE, sorted by Test AUC (exprs: expression data; clinical: clinical data; combined: expression and clinical data; net_exprs: expression only for proteins from the backbone network; net_combined: expression only for proteins from the backbone network combined with clinical/demographic data; fs: forward selection approach; lasso: LASSO selection).

| Combinations | lasso | fs | Validation (Mean AUC) | Test (AUC) |
| --- | --- | --- | --- | --- |
| net_exprs (GATv2Conv) |  |  | 0.83 | 0.82 |
| net_exprs (GATv2Conv) |  | ✓ | 0.85 | 0.81 |
| net_exprs (GATv2Conv) | ✓ | ✓ | 0.85 | 0.81 |
| net_exprs (GCNConv) |  |  | 0.84 | 0.81 |
| net_combined (GATv2Conv) |  |  | 0.90 | 0.80 |
| net_combined (GCNConv) |  |  | 0.90 | 0.78 |
| net_exprs (GCNConv) |  | ✓ | 0.83 | 0.78 |
| net_exprs (GCNConv) | ✓ | ✓ | 0.83 | 0.78 |
| net_combined (other ML) |  |  | 0.89 | 0.77 |
| net_exprs (other ML) |  |  | 0.82 | 0.77 |
| net_exprs (other ML) |  | ✓ | 0.82 | 0.77 |
| net_exprs (other ML) | ✓ | ✓ | 0.82 | 0.77 |
| exprs (other ML) |  |  | 0.89 | 0.76 |
| combined (other ML) |  |  | 0.91 | 0.74 |
| net_combined (other ML) | ✓ | ✓ | 0.89 | 0.73 |
| clinical (other ML) | ✓ | ✓ | 0.88 | 0.73 |
| clinical (other ML) | ✓ |  | 0.88 | 0.73 |
| net_combined (other ML) | ✓ |  | 0.89 | 0.72 |
| clinical (other ML) |  |  | 0.88 | 0.72 |
| combined (other ML) | ✓ |  | 0.97 | 0.71 |
| net_combined (other ML) |  | ✓ | 0.89 | 0.70 |
| exprs (other ML) | ✓ |  | 0.97 | 0.69 |
| combined (other ML) |  | ✓ | 0.95 | 0.69 |
| combined (other ML) | ✓ | ✓ | 0.98 | 0.68 |
| exprs (other ML) | ✓ | ✓ | 0.97 | 0.68 |
| exprs (other ML) |  | ✓ | 0.94 | 0.66 |
| net_exprs (other ML) | ✓ |  | 0.83 | 0.65 |

### Comparison of ML tools for Response to Treatment prediction

Using the EMMY data, we proceeded to perform a prediction with response to treatment, specifically for the drug empagliflozin. A similar 5-fold nested cross validation approach, as described above, was performed. Using regression ML model metrics RMSE and MAE, we were able to assess the performance of our models in predicting the value of NT-proBNP in visit 4 (26 weeks after first drug administration) using the correspond values from the previous 3 patient visits in combination with the available clinical data. The best performing combinations based on validation RMSE were obtained using LASSO-selected as well as LASSO and FS-selected features from visits 1, 2 and 3 with validation RMSE 0.46 and test RMSE: 0.59. Moreover, using only NTproBNP measurements from Visits 1, 2 and 3 resulted in the best generalization performance, with RMSE of 0.56 and MAE of 0.41 (see Table 7, Figure 8).

**Table 7.** Combination of features vs. classification performance RMSE and MAE for EMMY data prediction of NT-proBNP in visit 4, sorted by Test RMSE (v1: data from visit 1; v2: data from visit 2; v3: data from visit 3; ntprobnp: only NTproBNP data from corresponding visits; fs: forward selection approach; lasso: LASSO selection).

| Combinations | lasso | fs | RMSE_Validation | RMSE_Test | MAE_Validation | MAE_Test |
| --- | --- | --- | --- | --- | --- | --- |
| ntprobnp_v1_v2_v3 |  |  | 0.53 | 0.56 | 0.39 | 0.41 |
| v1_v2_v3 |  |  | 0.55 | 0.57 | 0.41 | 0.43 |
| v1_v2_v3 | ✓ |  | 0.46 | 0.59 | 0.36 | 0.45 |
| v1_v2_v3 | ✓ | ✓ | 0.46 | 0.59 | 0.36 | 0.45 |
| v1_v2_v3 |  | ✓ | 0.49 | 0.60 | 0.37 | 0.45 |
| v1_v2 |  |  | 0.60 | 0.63 | 0.45 | 0.48 |
| v1_v2 | ✓ |  | 0.52 | 0.65 | 0.40 | 0.51 |
| v1_v2 |  | ✓ | 0.54 | 0.65 | 0.42 | 0.51 |
| v1_v2 | ✓ | ✓ | 0.51 | 0.66 | 0.39 | 0.51 |
| ntprobnp_v1_v2 |  |  | 0.63 | 0.68 | 0.46 | 0.49 |
| v1 |  |  | 0.69 | 0.73 | 0.54 | 0.57 |
| v1 | ✓ |  | 0.65 | 0.75 | 0.51 | 0.59 |
| v1 | ✓ | ✓ | 0.64 | 0.78 | 0.51 | 0.62 |
| v1 |  | ✓ | 0.63 | 0.79 | 0.50 | 0.64 |
| ntprobnp_v1 |  |  | 0.87 | 0.90 | 0.67 | 0.70 |

**Figure 8.**
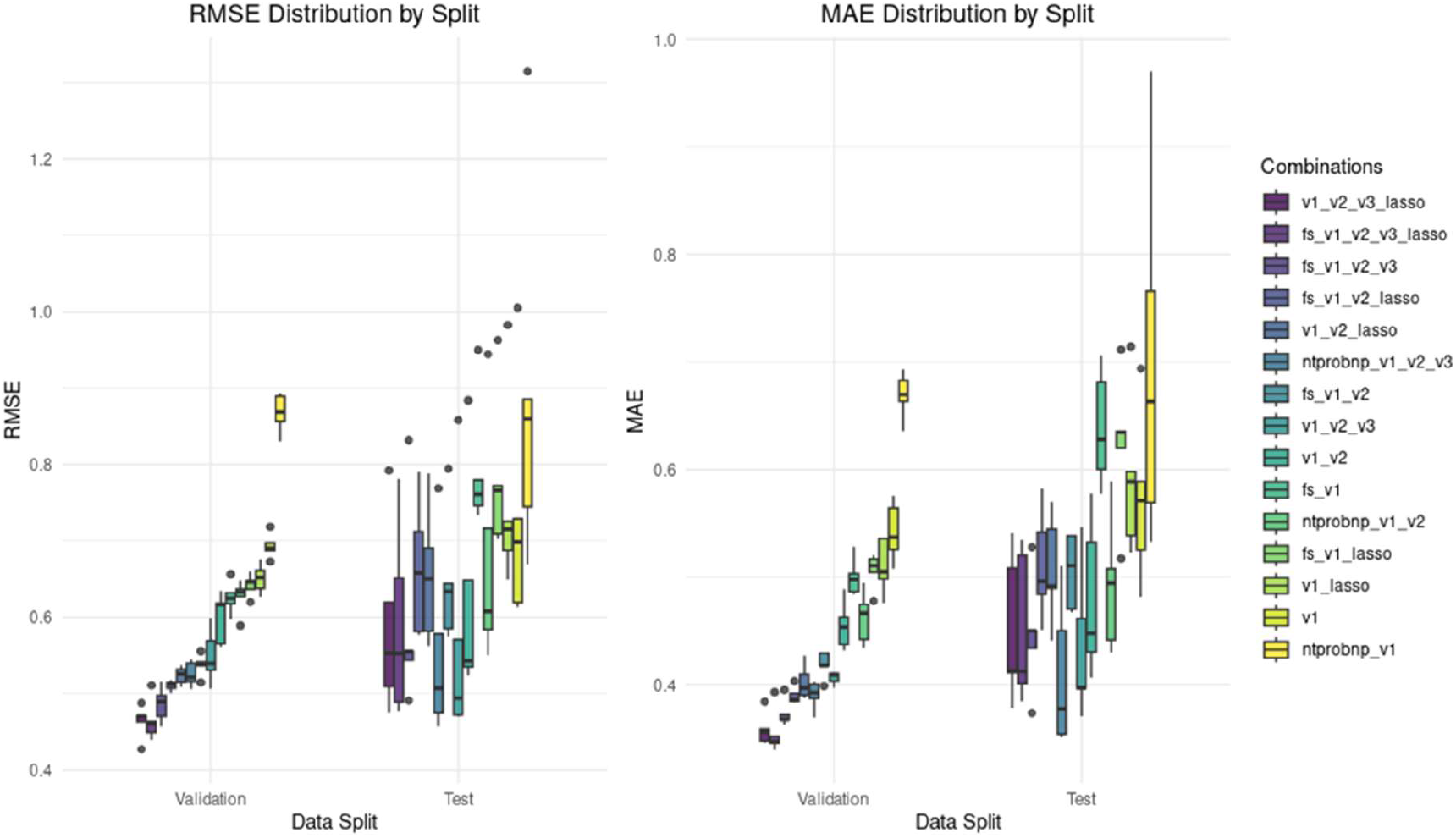
EMMY data top ML performance box plot displaying average over 5-fold nested cross-validation runs. Results were achieved using different ML models trained for prediction of NT-proBNP value at visit 4 which acts as the primary outcome change for the treatment group vs. the placebo group in the EMMY trial.

### Comparison of GNN and other ML models with Risk prediction Equations

It is common practice when analyzing CVD clinical trial data to apply certain risk equations that generally utilize a weighted sum of different clinical parameters to obtain a score and identify those individuals that are at higher risk for a CVD-related event. There are numerous such risk equations that provide clinical risk stratification such as the: GRACE score (Global Registry of Acute Coronary Events score), Framingham Risk Score (FRS), SCORE2 (European Society of Cardiology), QRISK3 (UK), Pooled Cohort Equations (American College of Cardiology / AHA), LIFE-CVD2 equation [31–36]. However, although the available clinical parameters were incomplete in order to perform the calculations from scratch in CDCS, IMMACULATE or EMMY datasets, the IMMACULATE dataset already included pre-calculated the GRACE score for the most patients. Therefore, we used only the samples with available GRACE score to calculate the AUC and compared it with our best-performing models, selected based on validation AUC (see Supplementary Table S3). For the GNNs, we used the best performing combination that can generate network. We observed that all of our models outperformed the GRACE score (see Table 8).

**Table 8.**
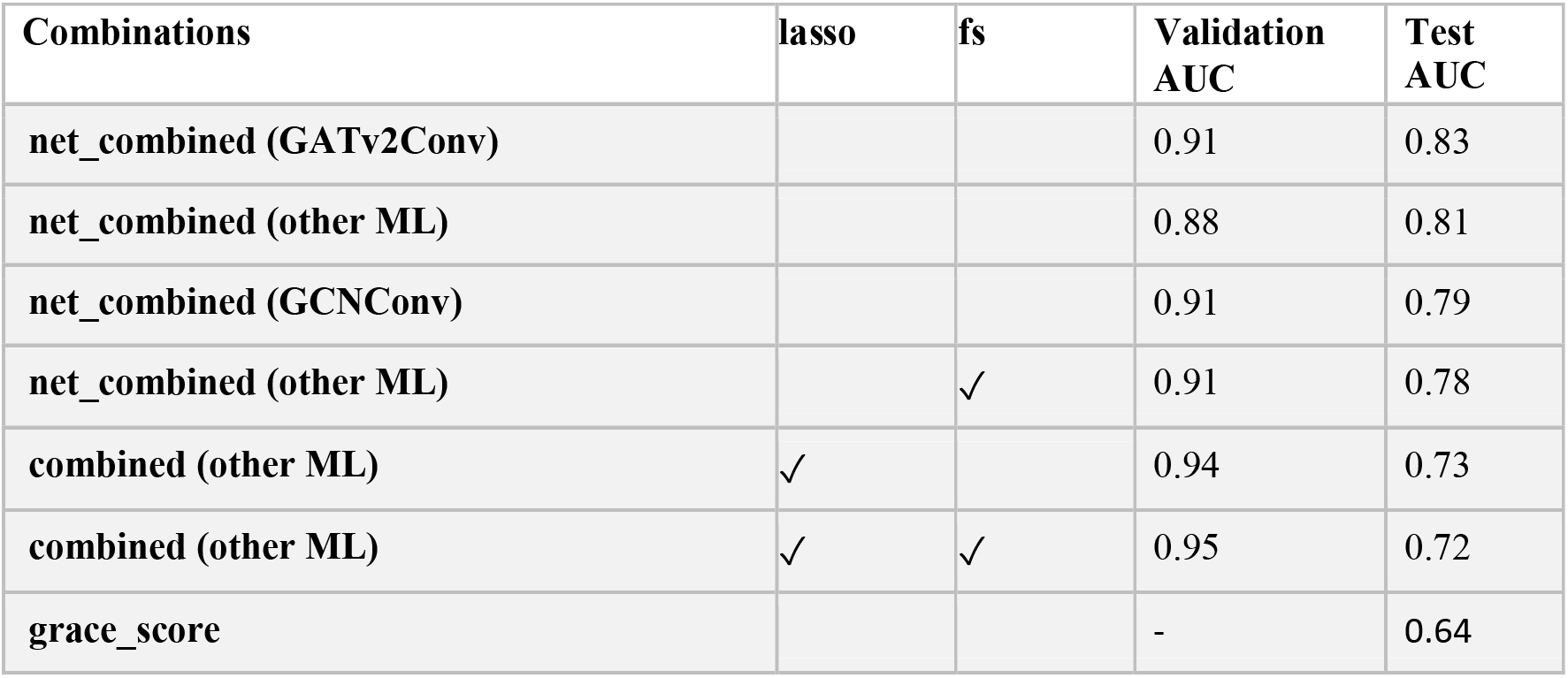
Comparison of model performance and AUC derived from the GRACE score with mean AUC values for validation and test sets across all evaluated models, sorted by Test AUC. (combined: expression and clinical data; net_combined: expression only for proteins from the backbone network combined with clinical/demographic data; fs: forward selection approach; lasso: LASSO selection) Note: immaculate_net_combined with FS-selected features was excluded from GNN models because the selected nodes formed a disconnected network.

### Kaplan Meier (KM) Feature Importance

We further assessed clinical feature importance using the Kaplan-Meier statistical technique. Although traditionally used to estimate survival probabilities over time, it can be particularly useful to analyze time-to-event data. Thus, we utilized the First_HF feature, which denotes the time in days before the HF event. We further grouped samples using additional *yes or no* features such as Beta blocker discharge, history of MI (hx_MI), history of HF (hx_HF), etc. Clear significance is shown for numerous of these features further enhancing feature importance estimations from our ML analysis (see Figure 9). Moreover, these features were among the most important clinical features selected by LASSO (see Figure 6B).

**Figure 9.**
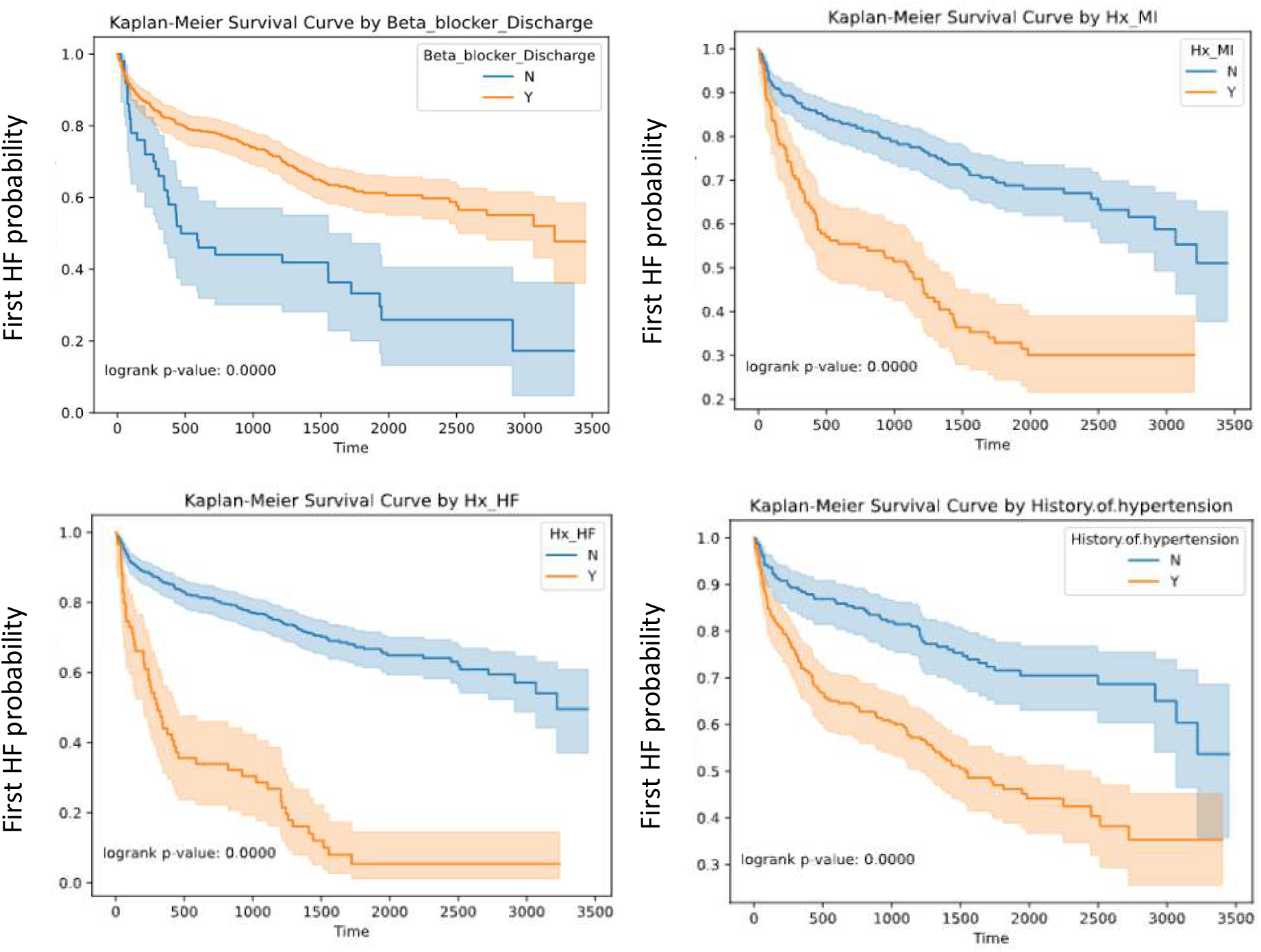
Kaplan-Meier statistical analysis for time-to-event data. Specifically, First_HF (days) was used in conjunction with multiple other clinical feature. Top 4 most significant features are shown (A, B, C and D).

## Discussion

Acute myocardial infarction (MI) remains a leading cause of subsequent heart failure (HF), yet the predictive landscape for patients who are at the extremely high-risk end of the spectrum of post-MI HF continues to face significant gaps. Despite decades of research identifying candidate biomarkers, the translation of these discoveries into routine clinical practice has been limited. Our study sought to address this limitation by integrating systems bioinformatics (SB) and advanced machine learning approaches, including Graph Neural Networks (GNNs), to improve both the identification of prognostic biomarkers and the computational stratification of patients at extremely high-risk of HF following MI.

Using prior knowledge and SB tools, like GeneMANIA, we first constructed a backbone cardiovascular network based on integrating multiple levels of information and knowledge. This network incorporated well-established CVD biomarkers—NT-proBNP/NPPB, TnT/TNNT2, CRP, and HbA1— along with genes containing well-characterized genetic variants associated with cardiovascular disease. Networks from STRING database were used with existing proteins of the dataset or subsets of the previously mentioned biomarkers. Using two large-scale, publicly available post-MI proteomic cohorts we then proceeded to overlay patient-specific proteomic and clinical data onto these networks in combination with additional prior knowledge. This enabled the generation of personalized patient networks, in which nodes represented biomarker expression levels, GDA, GDA-weighted expression levels, GO term – gene mapping and protein identity as well as edges captured similarity between the nodes. This framework effectively transformed heterogeneous, multi-source data into a unified integrated graph representation suitable for downstream machine learning.

Given the nature of our input (personalized patient networks) we proceeded with GNNs to obtain trained models for HF prediction post-MI. However, we also utilized other ML approaches for purposes of comparison. Results demonstrate that GNN models, particularly those employing attention mechanisms, outperformed traditional machine learning approaches (e.g., GLMs, XGBoost) on the cases with same data as well as GRACE score in predicting HF post-MI. Moreover, GNN models using only selected biomarkers, either alone or combined with clinical/demographic data achieved the best generalization performance on the external test dataset across all tested combinations. Attention-based training allowed the model to assign differential weights to protein connections, highlighting features with higher diagnostic/prognostic relevance. Notably, features known to be associated with CVD, such as NPPB/NT-proBNP, TnI/TNNI3, TnT/TNNT2 and patient history of HF consistently emerged as top contributors across ensemble ML models, reinforcing their established clinical relevance. Moreover, Kaplan-Meier analysis of time-to-event data corroborated the significance of a subset of these features, thus providing an independent validation of the predictive insights derived from the ML models. In addition to well known biomarkers for CVD, our model interpretability analysis highlighted key novel proteins with existing bibliography that supports their potential association with CVD. These included leukotriene A4 hydrolase (LTA4H), previously reported as a risk factor for sudden cardiac death due to MI and furthermore, genetic variation in LTA4H has also been associated with MI [37,38].

The integration of prior knowledge with patient-specific molecular and clinical data represents a key strength of this study. Specifically, GNNs trained without incorporating prior knowledge, in the form of GDA and GO terms, performed less accurately when it comes to generalization across datasets (data not shown). By leveraging SB approaches, we were able to enhance the interpretability of machine learning predictions, offering a potential pathway toward explainable Artificial Intelligence (AI) in clinical decision-making. This is particularly critical in the context of HF post-MI, where early identification of high-risk patients can inform preventative interventions and optimize treatment strategies.

The use of the EMMY trial data, where patients were treated with the drug Empagliflozin across visits, provides a unique setting and a longitudinal aspect that is not covered by the previous two proteomic datasets. Repeated patient visits and history together with laboratory results across time allow for a robust perspective on response to treatment and potential for timely intervention. High-risk individuals that received Empagliflozin appear to have significant changes in the primary outcome indicator (NT-ProBNP) with respect to the placebo group. Our regression model’s ability to predict NT-ProBNP levels at visit 4 (26 weeks after first drug administration) using longitudinal data, biomarker measurements, and clinical features could potentially enable timely therapeutic interventions and provide a secondary risk assessment ahead of critical time points.

Despite these promising findings, several limitations should be acknowledged. First, while our analysis incorporated three independent cohorts, external validation in larger, ethnically diverse populations is necessary to confirm generalizability. Second, the reliance on proteomic, molecular and clinical data alone may omit additional predictive layers, such as transcriptomic or metabolomic signals, which could further refine risk stratification. Finally, while GNNs offer powerful modeling capabilities, their computational complexity may pose challenges for real-time deployment at the point-of-care, emphasizing the need for continued efforts toward efficient, clinically deployable algorithms.

In conclusion, this study provides a proof-of-concept for integrated SB and GNN-based prediction of HF post-MI, highlighting both promising biomarker candidates and a scalable, explainable computational framework. In addition, regression ML analysis of longitudinal clinical and molecular data can substantially improve prognosis of high-risk individual and enable timely prediction of response to treatment, facilitating early therapeutic interventions. These results support the broader vision of Predictive, Preventive, and Personalized Medicine (PPPM), wherein multi-source data integration and advanced machine learning can facilitate early risk assessment, targeted intervention, and ultimately improved patient outcomes in cardiovascular disease.

## Data Availability

Data is available upon request

## Author Contributions

AO, NK and GMS conceived and designed the study and acquired data from HS and DL. AO and NK did the statistical analyses and developed, trained, and applied the convolutional neural networks. AO, NK, RA and TL implemented quality control of the algorithms. All authors interpreted the analyzed data and aided in conclusion inference. AO prepared the first draft of the manuscript. All authors contributed to manuscript revision and preparation.

## Data and Software Availability

The of proteomics dataset are available for download from the author’s GitHub page (https://github.com/ArisStefanosSn/HFproteomics). The EMMY trial data is available via data transfer agreement upon request. Full code for our GNN and regression models is available at: https://github.com/Bioinformatics-Department-C-BIG/CardioKGNN.

